# SEEG Contact Detector: A 3D Slicer Extension for Automated Localisation of Intracranial Electrode Contacts

**DOI:** 10.64898/2026.08.13.26360270

**Authors:** Jakub Smid, Petr Jezdik, Adam Kalina, Martin Kudr, Radek Janca

## Abstract

**Background:** Precise localisation of intracranial electrode contacts is essential for the interpretation of stereoelectroencephalography recordings and planning epilepsy surgery. In current clinical practice, this is typically a manual process, which is time-consuming and prone to variability. Existing automated solutions are often fragmented across multiple tools requiring technical expertise, limiting their adoption in routine clinical workflows. This study presents an open-source extension for 3D Slicer that provides an integrated, user-friendly standalone solution for the direct automatic detection of electrode contacts within a widely used medical imaging platform.

**Results:** The proposed method combines anchor bolt-based initialisation, probabilistic segmentation of electrode structures, and non-linear modelling to precisely track true electrode trajectories. The approach was evaluated on a dataset comprising 78 cases from 73 patients, including 1,078 electrodes with 14,480 contacts. The method achieved high localisation accuracy, with a median (interquartile range) deviation of 0.10 (0.06, 0.15) mm. Only 7/1078 (0.65%) electrodes required manual correction; these specific cases were handled using tools provided within the proposed extension.

**Conclusions:** The presented extension enables fast, accurate, and reproducible electrode contact localisation within a single integrated environment. By combining automation with intuitive user interaction, it significantly reduces processing time while maintaining clinical reliability. The tool’s free availability as an extension in 3D Slicer lowers the barrier to adoption and supports the standardisation of workflows across clinical and research centres.

## 1 Background

Stereoelectroencephalography (SEEG) is an invasive diagnostic technique used in epilepsy surgery, primarily for the identification of epileptogenic tissue, mapping of epileptic networks, and radiofrequency thermocoagulation in patients with drug-resistant focal epilepsy [2]. Precise knowledge of the anatomical positions of individual electrode contacts is thus crucial for the correct interpretation of SEEG recordings, stimulation mapping, and the planning of subsequent intervention and resective surgery [6, 9]. Traditionally, localising these contacts is a manual process in which clinical experts visually inspect post-implant imaging data (computed tomography, CT; and magnetic resonance imaging, MRI) [10]. More complex cases require a denser or more extensive coverage, involving a higher number of implanted multi-channel electrodes [1, 8]. While possible, the manual labelling of hundreds of recording contacts implanted in a single patient is exceptionally time-consuming, and can take several hours [2, 4]. As well as being time-inefficient, manual localisation is highly prone to human error, subjectivity, and inter-operator variability, which can compromise SEEG interpretation for surgical planning [2]. Furthermore, artefacts in the imaging data – particularly those arising from non-planar trajectories or converging electrodes – frequently render manual segmentations inaccurate or incomplete [6, 10]. These factors result in imprecise manual labels in three-dimensional (3D) coordinates; while such labels may be sufficient for visual navigation, they can lead to issues in subsequent processing, such as coordinate transformation to normalised anatomical Montreal Neurological Institute (MNI) space.

To address these profound limitations, the research community has developed numerous automated and semi-automated computational tools to extract electrode coordinates using post-implantation CT scans and pre-implantation MRI. For example, software suites such as EpiTools and its GARDEL module allow for the automatic segmentation, localisation, and anatomical labelling of SEEG contacts, significantly speeding up clinical interpretation [9]. Similarly, the iELVis toolbox provides an open-source pipeline that assists researchers in localising subdural electrodes and uniquely corrects for post-implant brain deformities such as brain shift [4], which can occur during craniotomy. Recent solutions like sEEG-Suite have integrated semi-automatic contact detection into larger multimodal platforms like Brainstorm [11] to support both reproducible research and complex clinical workflows [2]. Despite these computational advances, existing workflows remain problematic as they require users to engage with multiple disparate programs, programming languages, and commercial licenses [12]. Furthermore, while automated tools perform well under ideal conditions, they often fail to correctly parametrise semi-flexible electrodes that naturally curve or bend during physical insertion into brain tissue [6].

Consequently, there is a pressing clinical need for an integrated, easy-to-use tool that streamlines the localisation process within a single graphical user interface. This solution should be accessible to non-programmers and clinically oriented users in centres without appropriate technical support [12]. Integrating post-implantation processing tools directly into the free 3D Slicer platform [3] – as previously seen with the SEEG Assistant [10] – has proven highly effective in overcoming the limitations of complex command-line scripts. However, it still requires the input of preprocessed images from different software pipelines (co-registration, skull-stripping, segmentation). Specifically, the workflow proposed by Narizzano et al. [10] assumes the availability of both pre- and post-implantation CT scans. Before contact localisation, the pre-implantation CT must be affinely coregistered to the post-implantation CT, after which the two volumes are subtracted to remove the skull. The resulting image must then be manually thresholded to suppress the remaining brain tissue while preserving the metallic contacts. In addition to these preprocessing steps, the user must provide a Markups list containing the planned entry and target points of each electrode. Consequently, this proposed workflow does not provide an end-to-end solution. Furthermore, the software is not available through the 3D Slicer Extensions Manager, and requires users to complete several preprocessing steps and prepare the necessary inputs before contact localisation can be performed.

Building upon these aspects, we propose the *SEEG Contact Detector*, developed as an open-source 3D Slicer extension targeting clinical usability, the result of more than a decade of experience with SEEG at the Motol and Homolka epilepsy centres in Prague, Czech Republic and building on a previously published approach [6]. The developed extension provides an integrated end-to-end workflow that minimises manual overhead while offering robust graphical widgets for initialisation, visual inspection, and immediate result refinement. By implementing a detection pipeline that uses anchor bolt segmentation for initialisation of electrode segmentation combined with polynomial electrode tracing, this tool accurately models the true physical trajectories of implanted SEEG electrodes. This framework ultimately aims to equip clinics with an accessible, highly reliable solution to automate contact identification and to provide standardised, rapid SEEG localisation practices for the entire medical community.

## 2 Implementation

The SEEG Contact Detector was developed and evaluated as an open-source extension for 3D Slicer, accessible directly via the 3D Slicer Extension Manager (available from version 5.10.0). The extension is implemented as a scripted module written in Python and executed by the Python interpreter embedded in 3D Slicer. The design follows a user-friendly approach intended to streamline electrode localisation while providing intuitive tools for workflow initialisation, the visual inspection of results, and manual refinement of contact detections when required.

The extension consists of *Logical Class* (a computational engine containing the mathematical core, including image segmentation, axis estimation, and curve-fitting algorithms), and *Widget class* (the user interface layer, managing the graphical user interface – GUI – and user-driven interactions). The GUI is implemented using Qt Designer. The main algorithm blocks are summarised in the scheme in Fig. 1A; their functionalities will be described in detail in the following subsections. A module interface in the 3D Slicer is shown in Fig. 1B.

**Fig. 1.**
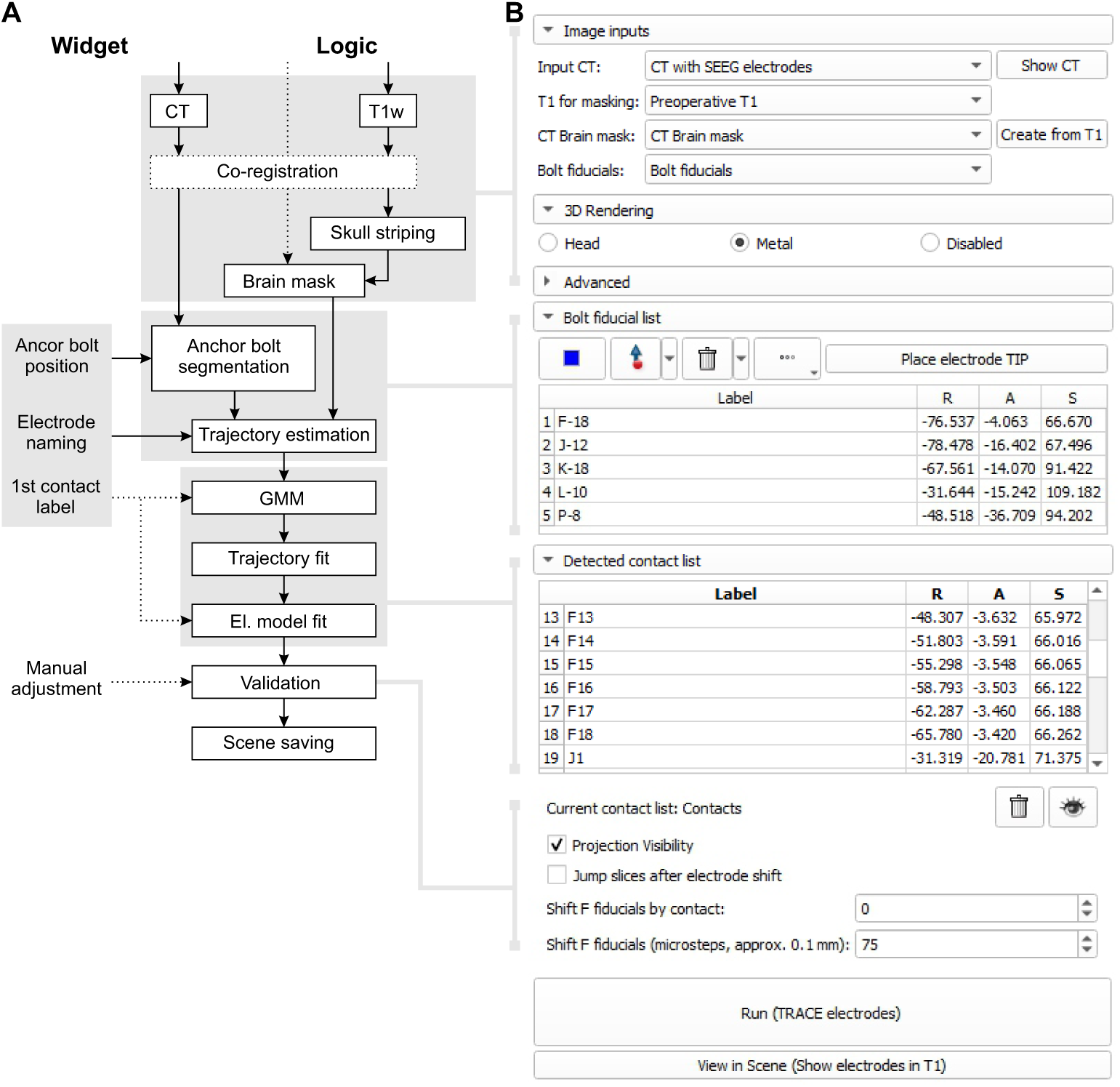
Scheme of the SEEG Contact Detector module. (A) Block scheme of the algorithm consisting of Widget and Logical classes. The dotted line represents optional user inputs. (B) The GUI of the SEEG Contact Detector module in 3D Slicer. The interface guides users through input selection, brain masking, anchor bolt segmentation, contact detection, and visual verification with optional manual correction. GUI: graphical user interface; SEEG: stereoelectroencephalography.

Post-implantation CT scans with metallic SEEG electrodes are the reference images for detection. MRI (T1-weighted recommended) is a complementary modality for determining intracranial space and anatomical background for the final multimodal view. Both modalities are rigidly co-registered.

Electrode wiring and connectors, temporarily placed beneath the sterile dressing after surgery, usually represent a source of metallic artefacts that distort and prohibit contact detection. Skull-stripping of the MRI isolates the intracranial space containing SEEG contacts (the recording sections of the electrodes), limiting the region to be detected; this process is detailed in section 2.2. Each electrode must be initially labelled and named by the user, placing a fiducial onto its anchor bolt to track the electrode’s trajectory. The label name is in the form of `prefix-suffix`, where the suffix is the number of contacts of the electrode, and thus also defines the physical length of its recording section. Labelled anchor bolts are segmented using the CT image and linearly fitted to a rough approximation of the electrode trajectory; this is detailed in section 2.3. However, in real-world conditions, semi-flexible electrode placement can deviate and curve from a straight trajectory, and implantation depth is not defined by default use.

Metallic intracranial objects (the contacts) are therefore assigned to individual electrodes; this is detailed in section 2.4. For each electrode, the metallic voxels are polynomially fitted in 3D space to estimate the real electrode trajectory. The contacts along the real trajectory are not detected individually, but as a group (a model of the recording section of the electrode) placed along the trajectory with defined spacing. The model is iteratively translated along the trajectory and correlated with the CT image to find the optimal position of the recording section; this contact detection is detailed in section 2.4.1.

The verification of all detections is accessible using the GUI. When necessary, the user can manually shift the whole group of contacts along the estimated real trajectory, for example, if the first contact is missed due to an X-ray shadow; this is detailed in section 2.5.

### 2.1 Data

We retrospectively collected data (CT and T1-weighted MRI scans) from 73 patients with pharmacoresistant focal epilepsy, who had undergone 78 implantations with SEEG electrodes (DIXI Medical MICRODEEP^®^) between 2017 and 2025. The dataset included three patients who required one re-implantation and one patient who required two re-implantations. SEEG exploration contained 13.8 ± 3.3 implanted electrodes with 185.6 ± 48.1 contacts on average. In total, the dataset contained MRI and post-implantation scans of 1,078 implanted electrodes with 14,480 electrode contacts. The distributions of electrodes and contacts across the dataset are shown in Fig. 2.

**Fig. 2.**
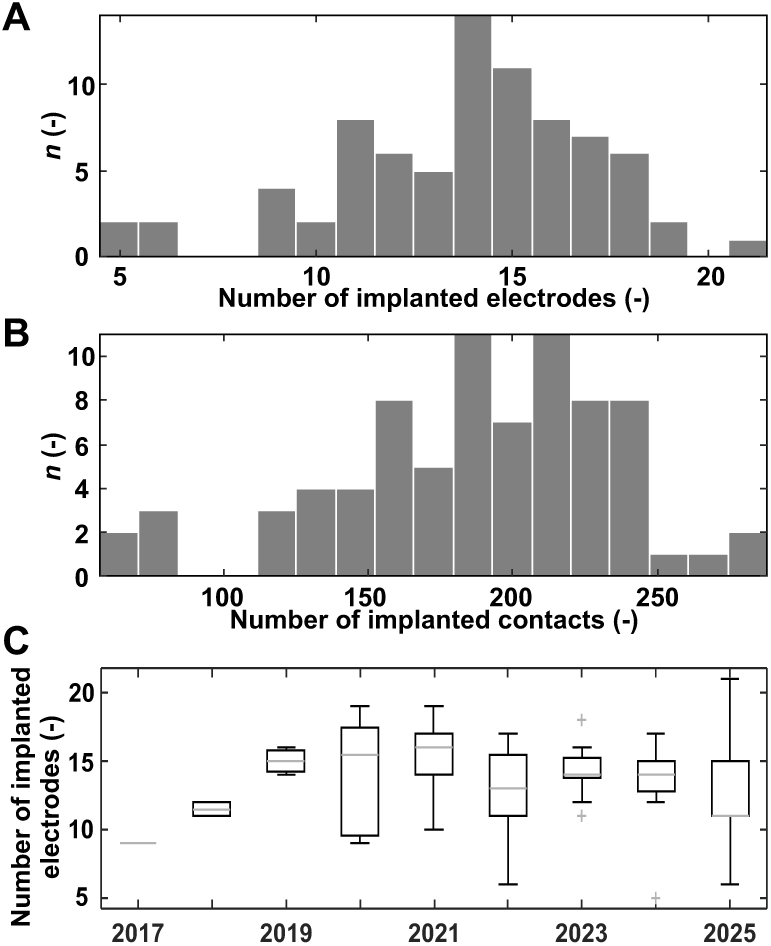
Characteristics of implanted electrodes. (A) Number of implanted electrodes per surgery across the dataset. (B) Number of contacts per implantation. (C) Number of implanted electrodes per surgery in years.

The post-implantation CT images (Toshiba/Canon Aquilion ONE or PRIME) were reconstructed using a soft-tissue window to keep Hounsfield units (HU). Image resolution was no coarser than (0.54×0.54×1) mm, (512×512×190) voxels respectively, and was typically (0.429×0.429×0.300) mm, (512×512×534) voxels. The pre-implantation 3T T1-weighted MRI images (Siemens MAGNETOM Vida or Philips Ingenia) had sub-millimetre resolution, typically (0.42 × 0.42 × 0.75) mm, (512 × 512 × 224) voxels.

Ground truth (GT) contact positions were labelled by an experienced clinical engineer (RJ) as part of routine clinical SEEG monitoring using the previously published pipeline [6], and were verified by epileptologists.

#### 2.1.1 Data preparation

The retrospective patient images were available in co-registered form. To simulate variability in image alignment and in the process of registration verification, we applied random rigid transformations to the T1-weighted images. Each image was randomly translated by up to 1000 mm in each direction and rotated by up to 10*^◦^* around each axis.

The anchor bolt fiducials were not available in the GT labels. For automatic placement, the position of the last contact was shifted by 10 mm along the electrode direction vector to approximate the centre of a typical 20 mm anchor bolt. This estimation successfully placed the anchor bolt fiducial outside the brain mask (within the anchor bolt area) for 1,019 of the 1,078 implanted electrodes. The anchor bolt positions for the remaining 59 electrodes were marked manually in the 3D Slicer viewer.

### 2.2 Image registration and skull-stripping

The post-implantation CT scan in a soft-tissue reconstruction window is the main referential volume for electrode detection. The complementary T1-weighted MRI is used as anatomical background and input for skull-stripping to obtain the intracranial region of interest, ROI_brain_. For this purpose, both images must be rigidly co-registered.

In practice, the direct rigid image registration of moving MRI to fixed CT often failed due to the amount of high-intensity metallic voxels in the CT. To resolve this, the CT was thresholded to remove metallic parts, keeping bone and soft tissue only (HU *<* 3000). This resulted in successful registration in all cases. For this step, the extension uses an embedded module of the BRAINS algorithm [7] with the following settings: 1% of samples, Initialize Transform Mode – Geometry Align, Rigid registration (six degrees of freedom). The input volumes keep their original orientation, and the obtained registration transformation is applied for visual projection.

To eliminate interference from metallic parts (electrode wires, connectors) that are unimportant for contact detection and cause imaging artefacts, the CT is masked by ROI_brain_, automatically computed from T1-weighted MRI (default) using the HD-BET algorithm [5]; example in Fig. 3H. The obtained brain mask (binary) is reoriented to CT by the registration transform.

**Fig. 3.**
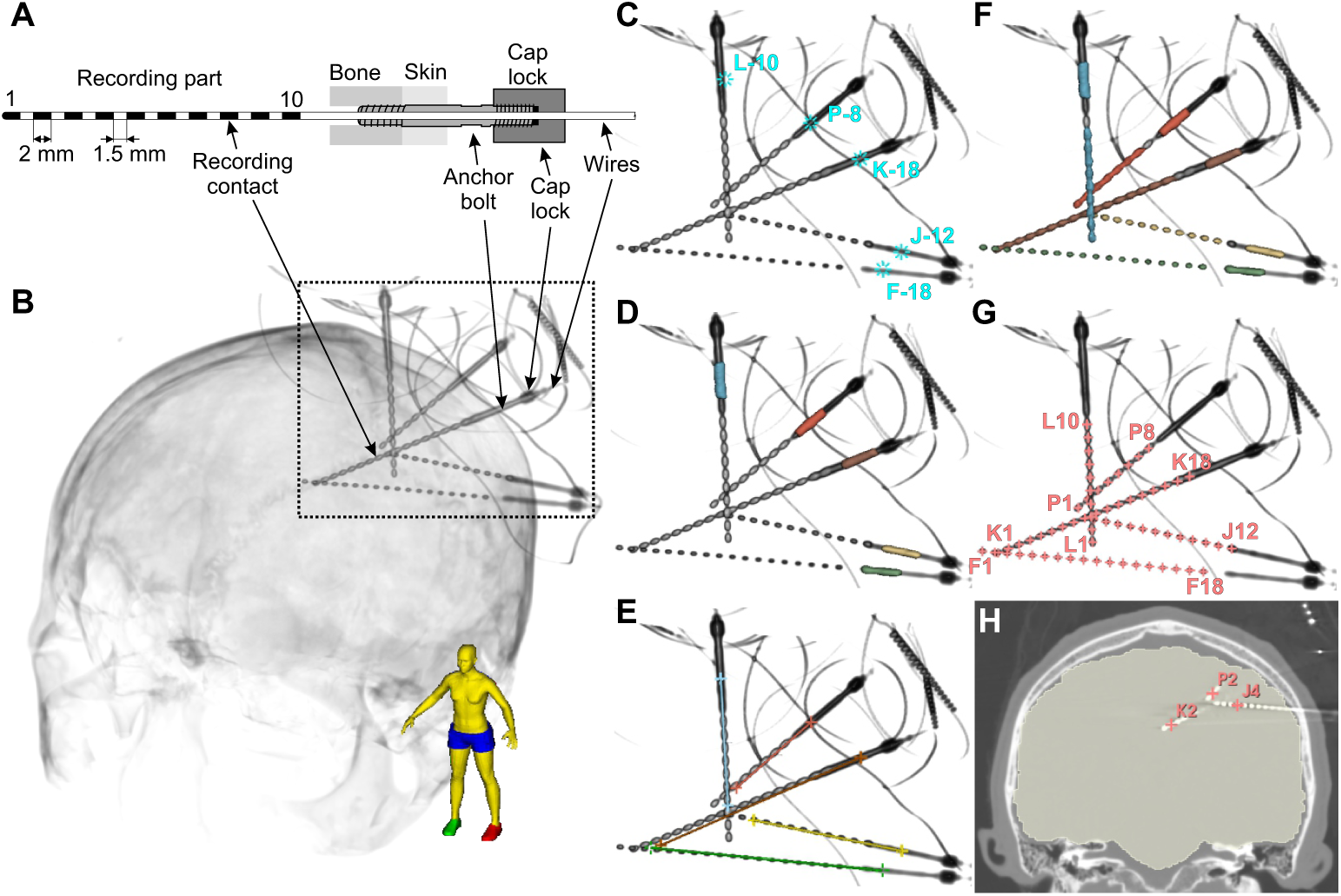
Overview of the SEEG Contact Detector processing pipeline. (A) Scheme of a SEEG electrode with recording contacts, anchor bolt, and locking cap. (B) 3D visualisation of the post-implantation CT, showing the skull with implanted electrodes. Dotted rectangle marks region of detail. (C) Fiducial placement, with electrodes named according to their number of contacts. (D) Anchor bolt segmentation from a post-implantation CT scan. Coloured regions represent individual segmented anchor bolts used to estimate initial electrode trajectories. (E) Anchor bolt axis estimation. A segmented bolt provides an estimate of the electrode axis used for GMM initialisation. (F) Electrode segmentation. Metallic voxels within the brain region are assigned to individual electrodes using a GMM, with each colour representing a separate electrode. (G) Polynomial estimation of electrode trajectory and electrode model fitting determine each contact position. (H) Visual inspection and editing of detected contacts can be performed in CT images. Semitransparent yellow represents ROI_brain_. SEEG: stereoelectroencephalography; CT: computed tomography; GMM: Gaussian mixture model.

In the GUI, the definition of input images, registration, and skull-stripping represents the first step of the SEEG Contact Detector pipeline. *Bolt fiducials* are defined in the next section, 2.3.

### 2.3 Anchor bolt segmentation

Anchor bolts (DIXI Medical MICRODEEP) have standardized sizes, ranging from 15 to 35 mm (20 and 25 mm used in our centre), with a 2.45 mm outer diameter (Fig. 3A–C). The first step of the algorithm is segmentation of the anchor bolts in the post-implantation CT scan. To initialise this step, the user provides approximate anchor bolt locations by placing fiducial points for all electrodes on the CT image in the GUI (Fig. 3C). The user can place the points directly in slice views (axial, coronal, or sagittal) or in the 3D rendering window. Fiducial points placed in the 3D view are automatically snapped to the first non-zero voxel intersected by the viewing ray ^1^. For this purpose, the user can switch between three options for 3D rendering in the GUI using the interface or keyboard shortcuts, allowing easy orientation and thresholding of the CT image to localise individual electrodes fixed in an anchor bolt: (F5) Head shows the skull and metallic electrodes; (F6) Metal shows metallic parts (HU *>* 3000) only; (F7) Disabled hides the 3D render. Anchor bolt fiducials should be placed in the 3D rendering window using the Metal rendering mode to ensure the fiducial snaps correctly onto the bolt’s surface.

Placed fiducials are stored in a *MarkupsNode*, which is a standard 3D Slicer data structure representing a list of user-defined points in “left to Right, posterior to Anterior, inferior to Superior” (RAS) coordinates. For optimal results, the fiducial should be placed in the middle of the anchor bolt or far away from other metallic parts.

Each fiducial must follow the `prefix-suffix` naming convention, where the prefix represents the alphanumeric electrode name and the suffix indicates the number of contacts of the corresponding electrode; the final symbol “-” in the character string is a separator. The placed fiducials appear in the *Bolt fiducial list* section of the module interface. Newly created fiducials are automatically assigned letters in alphabetical sequence, which can be edited by the user.

Once the fiducial positions are provided, a spherical (ROI_bolt_) is extracted around each anchor bolt location from the CT image. Restricting the segmentation to ROI_bolt_ helps prevent inclusion of other metallic structures, such as electrode wires or external connectors, that may be present near the scalp surface. The radius of this region is controlled by the user-tunable parameter *Radius around the bolt* (default: 5 mm). Within ROI_bolt_, metal structures are segmented using intensity thresholding with the parameter *Metal threshold* (default: 3000 HU). The threshold isolates high-intensity metallic voxels in the CT volume. In some cases, metal voxels from neighbouring anchor bolts may still appear within ROI_bolt_. Selecting the largest-connected-component analysis with 26-neighbour connectivity ensures that only the intended anchor bolt is retained (Fig. 3D).

#### 2.3.1 Anchor bolt axis estimation

The trajectory of each electrode passed through the bolt is estimated from the segmented anchor bolt obtained in the previous step. In addition, a brain mask in the CT image space is required to determine the direction of electrode insertion (from surface to intracranial space).

First, the direction of the electrode is estimated using principal component analysis (PCA) applied to the voxel coordinates of the segmented anchor bolt. This provides both the centroid of the bolt segmentation and a vector corresponding to the dominant direction of the bolt. This vector serves as an initial estimate of the electrode trajectory. The manually placed anchor bolt fiducial is orthogonally projected onto the electrode trajectory; this projection is taken as the entry point (EP) of the electrode at the patient’s head. The electrode tip (TP) is roughly estimated by extending the trajectory from EP into the intracranial space by the physical length of the electrode *L*, computed by equation 1.

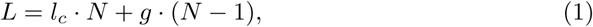

where *l_c_* represents the contact length, *g* is the gap distance between contacts, and *N* is the number of contacts of the electrode. *N* is extracted from the label of the corresponding anchor bolt fiducial. The default values of the DIXI Medical SEEG electrodes are *l_c_* = 2 mm and *g* = 1.5 mm.

However, the axial direction from EP into the intracranial space is not defined. Therefore, the TP is placed in both directions, and the one located within the ROI_brain_ is selected.

The TP and EP points define the initial estimate of the electrode placement for electrode contact tracing. An example of the resulting trajectory estimation is shown in Fig. 3E.

### 2.4 Clustering of electrodes

The goal of this step is to identify all metallic voxels belonging to SEEG electrodes in the post-implantation CT scan and assign them to individual electrodes.

Before clustering, a voxel array is constructed that contains only data relevant for the detection task: voxels in CT ∈ (ROI_brain_ ⋃ ROI_bolts_) and HU *>* 3000. These voxels correspond to metallic structures, such as electrode contacts and anchor bolts.

To assign metal voxels to individual electrodes, we apply a Gaussian mixture model (GMM), in which each electrode is represented by a single Gaussian component. Proper initialisation of the GMM is critical for accurate clustering. For each electrode, the initial covariance ellipsoid is aligned with the EP–TP line segment, whose direction defines the major principal axis. To regularise the covariance matrix, an isotropic variance (1% of the maximum variance along any single axis) is added to all three dimensions. All Gaussian components are initialised with equal weights. The subsequent GMM optimisation only refines the parameters of the initialised ellipsoids to better fit the contact voxels corresponding to each electrode’s real placement (Fig. 3E–F).

The clustering of metallic voxels limits a region for the contact detection of individual electrodes, ROI_el_. An example of the resulting electrode clustering is shown in Fig. 3F.

#### 2.4.1 Electrode trajectory and contact model fitting

Due to imaging artefacts in CT, some contacts may contain voxels with low or negative HU-values at their centre or in regions of X-ray shadow, producing small voids in ROI_el_, primarily when using non-recommended bone-window reconstruction. Therefore, several iterations of the morphological closing operation (dilation followed by erosion) are applied. The number of iterations is determined based on the physical diameter of the electrode contacts and the spatial resolution of the CT scan:

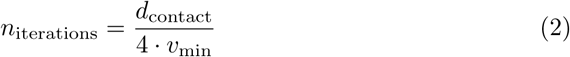

where *d*_contact_ represents the physical diameter of the contact (default: 0.8 mm) and *v*_min_ denotes the minimum physical voxel spacing of the input CT volume.

The fitting of the electrode trajectory in 3D space is formulated as three polynomial fits for the spatial coordinates: *x*(*s*), *y*(*s*), *z*(*s*). First, an initial linear trajectory is computed using PCA applied to the ROI_el_. The first principal component corresponds to the linear axis, *s*, along the electrode shaft, onto which the coordinates of the ROI_el_ are projected.

To model potential bending of the electrode inside the brain, the trajectory is refined by fitting three fifth-degree polynomials independently for each spatial dimension, parametrised by the axis *s*. Intensities of the CT volume are used as weights of the fitting algorithm. Finally, the continuous axis *s* is extrapolated beyond the last voxel by two contact lengths plus two inter-contact gaps (default: 7 mm), and the polynomials are discretely sampled with a resolution of 0.1 mm.

After estimating the trajectory, the centres of the electrode contacts are determined using a physical electrode model. The electrode model is represented by a sequence of Gaussian-shaped elements (“Gaussian spheres”) corresponding to individual contacts. The standard deviation in each axis is defined by the contact diameter *σ_mm_*, which controls the spatial extent of the Gaussian. The magnitude of the elements is 1 at its centre, as the normalisation factor is omitted. The shape of the region modelled by the Gaussian sphere is controlled by the parameter *Gaussian blob size* (default: 3*σ_mm_*). The dimensions of the model reflect the known physical properties of the electrode, including contact length and spacing.

The electrode model is moved along the fitted trajectory, starting from the innermost point of the curve toward the EP. Once all hypothetical contacts of the model overlap with metal voxels, the algorithm performs cross-correlation between the electrode model and the CT image across translations corresponding to one contact length plus one inter-contact gap^2^. The position with the maximum correlation is selected as the most probable location of the electrode contacts.

The final detected contact centres correspond to the centres of the Gaussian elements of the best-fitting electrode model. An example of the fitted electrode model is shown in Fig. 3G–H.

### 2.5 Verification of contact detections

The contact positions obtained from the best-fitting electrode models are stored in a new Markup list named *Contacts*. Contact naming follows user defined labels of the corresponding anchor bolt, e.g., F-18: F1, F2, …, F18.

All detected contacts are displayed in the *Detected contact list* of the GUI, allowing users to quickly inspect individual detections. Selecting a contact from the list refocuses slice views (axial, sagittal, coronal) to its position.

Below the list, users can toggle the visibility of the detected contacts or remove the entire detection list if needed. Additionally, the projection visibility can be enabled or disabled. This option controls whether contacts are shown in slice views even when they are not located in the currently displayed slice.

#### 2.5.1 Troubleshooting and manual editing

During previous detector development and clinical practice [6], detection of the first contact tip occasionally failed in dense SEEG exploration with substantial X-ray artefacts around touching electrodes. This results in lower CT intensity of the first contact and better correlation of electrode model fit starting at the second contact.

All detected contacts of the selected electrode can be adjusted at once along the fitted electrode trajectory while still respecting the physical model of the electrode.

To shift the detections, the user must first select any contact of the electrode in the *Detected contact list*. Two options are available: *Shift fiducials by contact* and *Shift fiducials (microsteps)*. The first option shifts the contacts of the selected electrode by the length of one contact length and one inter-contact gap. Using this control, the electrode can be moved up to two contacts toward the EP or toward the electrode tip.

Alternatively, the *microsteps* option allows finer adjustments along the fitted trajectory. The specified value corresponds to the number of discrete positions along the fitted curve. Shifting the electrode by one microstep therefore moves the contacts by one sampling point on the fitted trajectory (approximately 0.1 mm).

The module also allows manual placement of the electrode tip during the anchor bolt labelling described in section 2.3. The tip can be defined as a fiducial within the *Bolt fiducial list*. This fiducial must follow the naming convention prefix-1 and should be placed at the tip of the first contact blob in metal rendering.

When the detection algorithm is executed, this manually placed point is used to initialise the GMM and defines the starting position for the correlation-based model fitting. The best model fit is then searched within a distance corresponding to one contact length plus one inter-contact gap from the placed point. Providing the electrode tip in this way bypasses the requirement for all electrode voxels to be detected as metal during the initialisation step (for low-quality CT).

The rarest failure occurred in cases with previous craniotomy, leaving objects such as metal bone clips or shunts. In these cases, when brain mask segmentation includes metallic parts within ROI_brain_ that are not from SEEG electrodes, the GMM algorithm assigns these voxels to a random electrode in their axes. Physical electrode trajectory and model fitting is therefore distorted, resulting in a strongly deviated trajectory or a missing first contact. However, the 3D Slicer allows mask editing by default, and manual editing of ROI_brain_ solves this failure.

### 2.6 Saving and clinical visualisation

The results of the processing blocks of the extension are, by default, automatically saved to the directory containing the CT image in file formats commonly used by 3D Slicer to protect against unexpected application crashes, although none were observed during development. The rigid transformation between CT and MRI is stored as `transform_T1_to_CT autosave.h5`, brain mask as `CT_brain_mask_autosave.seg.nrrd`, the list of anchor bolt fiducials as bolt fiducials autosave.mrk.json, and the list of detected contacts as contacts_autosave.mrk.json.

Once the user has finalised and verified the detections, clicking the *View in Scene (Show electrode in T1)* button creates a scene for clinical inspection, see Fig. 4. All image modalities are reoriented to the native T1-weighted image space, which is used as the background and typically serves as the reference modality for other neuroimaging data and preoperative examinations. The CT lookup table is changed to the blue *Ocean* colour map, thresholded (HU *>* 3000) to enhance the visibility of metallic structures, and displayed as the foreground. The result can be saved by the user as either an MRML scene or a scene bundle for future use.

**Fig. 4.**
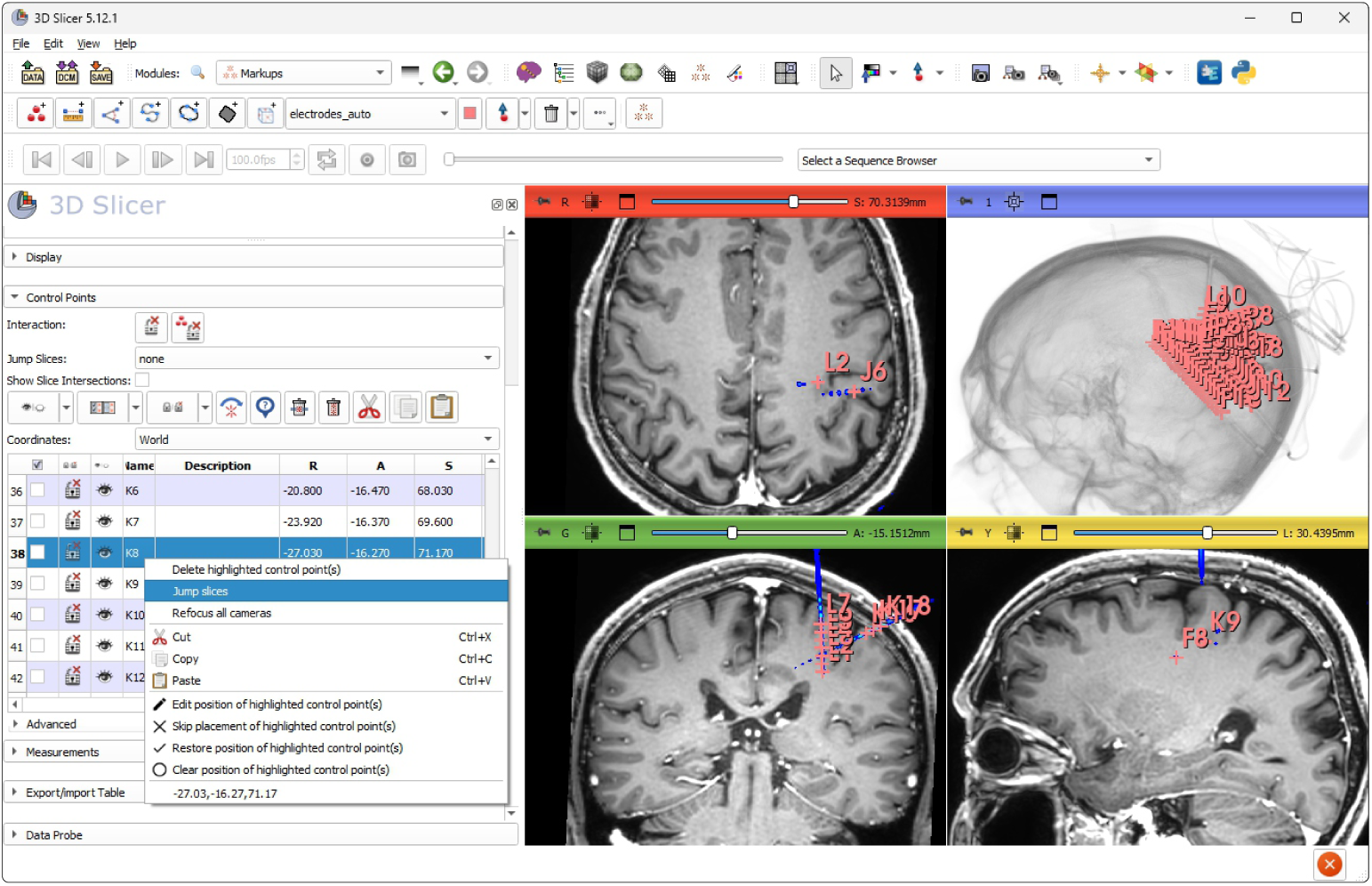
Final clinical view. Created scene composed of background T1-weighted image, co-registered CT with thresholded metalic SEEG electrodes in blue colour palette, and fiducials of detected contacts. The standard module Markups allows fast navigation by “Jump slices” function, which displays the selected contact in the axial, sagittal, and coronal views.

## 3 Results

The SEEG Contact Detector was implemented as a standalone extension for the 3D Slicer viewer, the core of which was derived from the original MATLAB implementation without an interactive GUI [6]. Outputs of the MATLAB implementation represented the GT reference against which the SEEG Contact Detector was verified. The verification was conducted in developer mode, which automatically loads and processes the prepared dataset as a single batch. This functionality is available in the *testing* branch on GitHub.

### 3.1 Detection failures required manual correction

Detection accuracy was evaluated using the Euclidean distance between GT positions and the corresponding positions estimated by the SEEG Contact Detector, hereafter referred to as the deviation. For each electrode, the mean deviation across all contacts was computed. Electrodes with a mean deviation exceeding 1 mm were considered failed automatic detections requiring visual inspection. Such failures occurred in only 7/1078 (0.65%) electrodes, concentrated in specific conditions related to patients’ individual cases (4/78; 5.1%), described below.

#### Touching at the electrode tip

The electrodes’ trajectories can cross in dense SEEG exploration, with the tip of one electrode potentially touching another (tip to tip or tip to middle section). The touching contacts are blended together in the CT image. The GMM can not assign the first contact (tip) to the correct electrode, and shifts all contacts by one position. This was solvable using one-click correction in the GUI. In clinical practice, this is the most common failure mode (4/7 failed electrodes).

#### Other metallic object in intracranial space

This type of failure (1/7 failed electrodes) occurred in a patient with a previous craniotomy and a residual cranial fixation plate. Metal-induced susceptibility artefacts in T1-weighted MRI result in imprecise extraction of the brain mask (ROI_brain_) using the HD-BET algorithm [5] and reduce the accuracy of CT–MRI registration through local B_0_ field distortions. Consequently, the registered brain mask erroneously overlapped with the cranial interlink plates in the post-implantation space. Occurrence of other non-electrode metallic voxels in the ROI_brain_ resulted in incorrect electrode trajectory fit and model placement, and thus contact detection errors. Manual editing of the ROI_brain_ mask to exclude interlink plates and subsequent re-running of the detection resulted in correct contact identification. In clinical practice, when a patient has undergone any neurosurgery procedures involving permanent implants (e.g., clips, interlink plates, or shunts), the brain masking results must be verified and manually cropped.

#### Touching anchor bolts

When anchor bolts were located very close to each other (touching or crossing), combined with inaccurate fiducial placement, the initial bolt segmentation and trajectory estimation failed during the initialisation of GMM. In dense SEEG exploration, GMM may not correctly assign individual electrodes, resulting in incorrect polynomial trajectory fit and final detection. The manual correction of anchor bolt fiducial position in two electrodes and subsequent re-running of the algorithm resulted in correct detection. In clinical practice, if anchor bolts are close or touching, precise fiducial placement is recommended, ideally positioned at the point of maximum separation from adjacent anchor bolts. Optionally, an additional fiducial at the electrode tip improves initialisation robustness in all scenarios.

#### Metal artefacts in CT

In one case, photon starvation and beam hardening artefacts degraded CT in the region around the electrode’s anchor bolt. The tip of one electrode had to be manually specified during anchor bolt segmentation. Additionally, some other contacts were not clearly visible in the CT image, and model fitting skipped the first contact; this was one-click shift corrected.

All failed detections were quickly resolved (within a few minutes) directly within 3D Slicer, using the tools provided by the module and accessible through the GUI. A video guide is available on the project page (section 5.2). After applying the described manual adjustments, the corrected contact positions were saved and included in the subsequent analysis of detection accuracy.

### 3.2 Accuracy of detection

We evaluated the deviation of detected contacts using the GT dataset. The median (interquartile range) was 0.10 (0.06, 0.15) mm, with a maximum of 1.07 mm. The median deviation was within voxel resolution. The distribution of contact deviations is shown in Fig. 5A.

**Fig. 5.**
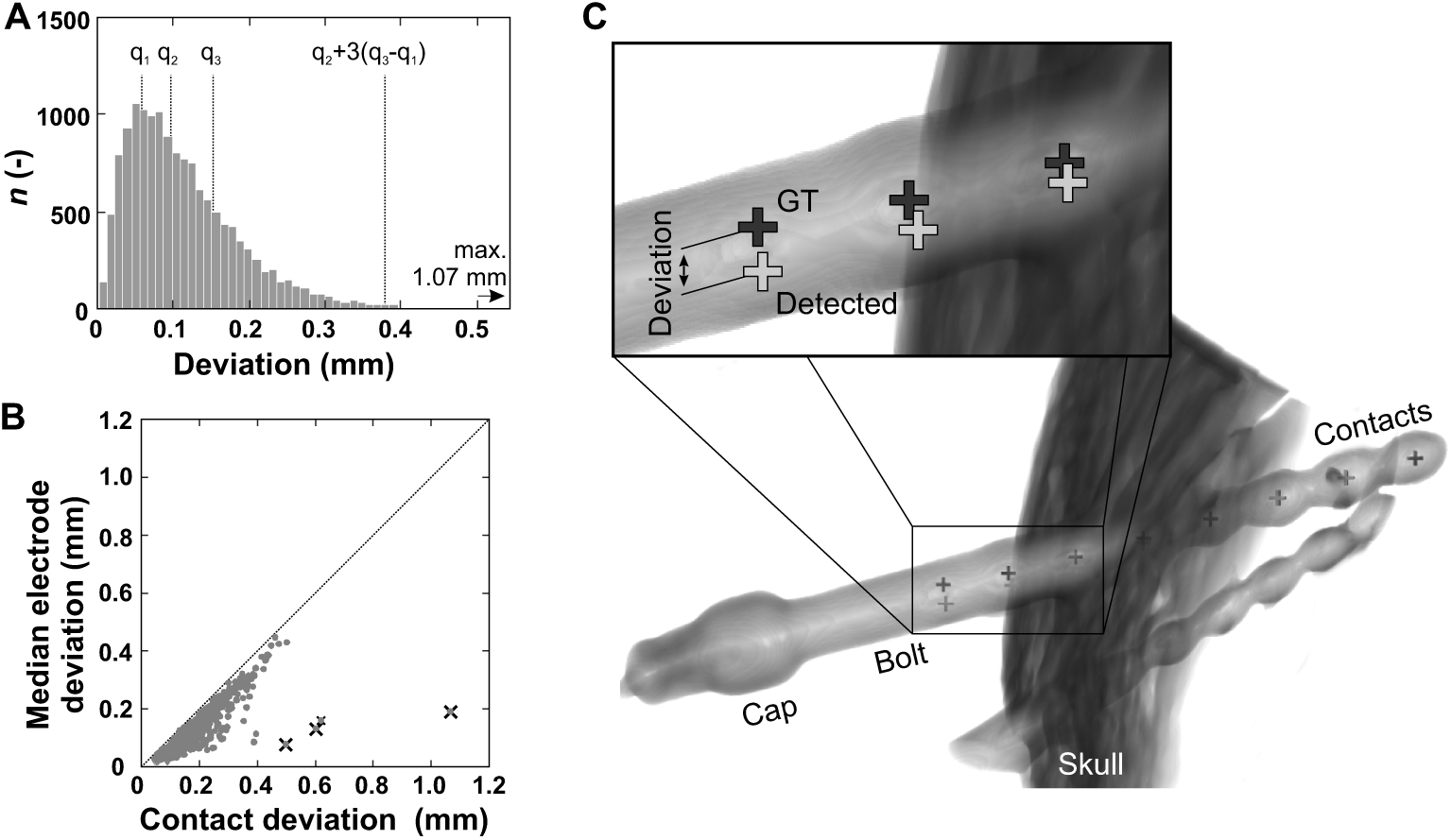
Comparison of detected contacts with reference GT. (A) Histogram of all contact deviations. (B) Maximal versus median contact deviation per electrode, revealing outlier detections, marked by (*×*). (C) Visual inspection of the most deviated detection reveals imprecise detection of contacts lying outside the brain in the anchor bolt hole. The 3D CT visualization of the implanted electrode shows imprecise trajectory fitting of contacts within the bolt, also for GT. CT: computed tomography; GT: ground truth.

To identify outliers, we compared instances in which a single contact exhibited a high deviation while the remaining contacts of the same electrode had low deviation. This produced a plot of the maximal versus median deviation for each electrode, as well as for each patient’s implantation (Fig. 5B). Visual inspection of the most significant outlier contacts (4/1078 electrodes across 3/78 implantations) revealed that these contacts were located inside the anchor bolt hole (typically the last contacts of the electrode); that is, these contacts were outside of the brain and therefore excluded from clinical processing (Fig. 5C). The position of the GT was also inaccurate in these cases.

### 3.3 Consistency of detection

To assess the consistency of the SEEG Contact Detector and identify potential systematic errors, we evaluated whether the detector exhibited bias in the RAS coordinate system^3^.

Electrode trajectories are planned to avoid transhemispheric trajectories whenever possible. While our implementation can process electrodes that cross both hemispheres, the dataset we used for testing contains only unilateral electrodes. Displacement in the median (interquartile range) form was –0.02 (–0.09, 0.02) mm from the centre to the left, and 0.03 (–0.01, 0.09) mm to the right. The minimal offset between left and right orientation was 0.08 mm, which may have been introduced by rounding in transformations between voxel and real-world (RAS) image spaces (Fig. 6A).

**Fig. 6.**
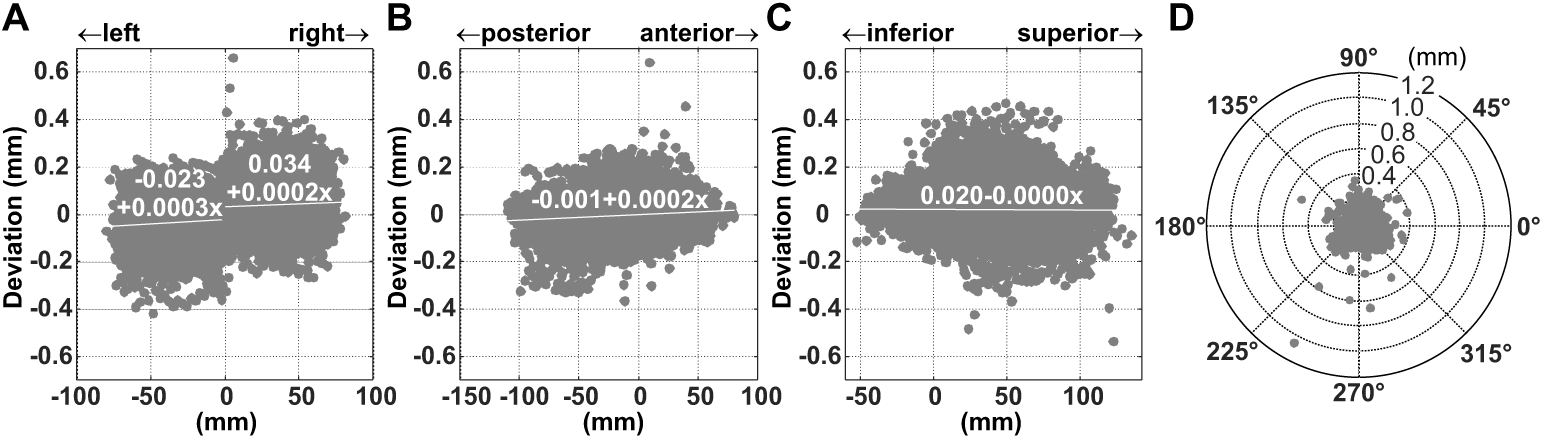
Detection consistency dependent on RAS contact position. Bland–Altman diagrams visualise the (A) left–right axis, (B) anterior–posterior axis, (C) inferior–superior axis. The linear regression (*y* = *a* + *bx*) shows minimal dependency on the position from the centre of the head. (D) Radial deviation visualised in the probe view (distance of orthogonal projection to electrode axis).

The effect of contact position on deviation was also minimal in the posterior-anterior –0.004 (–0.03, 0.02) mm and inferior-superior 0.01 (–0.01, 0.05) mm axes (Fig. 6B–C).

Radial deviation – the distance of contact detection from the orthogonal projection to the electrode axis – was 0.03 (0.02, 0.05) mm, demonstrating a stable axis fit (Fig. 6D).

We analysed the dependency of deviation on the order of contacts, reflecting their depth along the electrode. We assumed that cumulative error in the model’s contact displacements would grow, due to sampling of the polynomial trajectory fit. This assumption was not confirmed; inter-contact spacing for single electrodes remained stable.

All observed differences had no impact on the clinical interpretation, and therefore confirm detector reliability and reproducibility.

## 4 Conclusions

In this work, we introduce the SEEG Contact Detector, an extension for 3D Slicer designed to provide fast, accurate, and clinically accessible localisation of SEEG electrode contacts in a standalone application. By integrating the entire detection pipeline into a single GUI, the proposed solution addresses key limitations of existing workflows: namely, their fragmentation across multiple tools, reliance on programming expertise, and limited robustness in challenging clinical scenarios. By making the SEEG Contact Detector available in the 3D Slicer’s Extension Manager, we aim to promote standardised, reproducible, and efficient SEEG contact localisation across clinical and research centres.

The presented method combines anchor bolt-based initialisation, probabilistic electrode segmentation, and non-linear trajectory modelling to accurately capture the true geometry of implanted electrodes, including physiologically realistic bending. Quantitative evaluation on a large and diverse clinical dataset demonstrated high accuracy and robustness across a wide range of implantation configurations. Importantly, cases requiring manual intervention were rare, and could be efficiently resolved using intuitive tools directly within the 3D Slicer environment, without the need for external processing. The method achieved an accuracy at the subvoxel level.

From a clinical perspective, the main contribution of this work lies not only in the accuracy of the detection algorithm, but also in its usability and integration into an established imaging platform. The extension is designed to fit naturally into existing clinical workflows, reducing processing time from hours to minutes while maintaining transparency and user control. This combination of automation and interactivity is essential for adoption in routine practice, where reliability and ease of use are critical.

Implementation as an extension in the 3D Slicer environment allows future integration, for example of tissue segmentation, anatomical parcellation, and transformation to normalised MNI coordinates.

Overall, the proposed extension represents a practical step toward wider adoption of automated SEEG processing and has the potential to significantly streamline clinical workflows and improve consistency in epilepsy surgery planning.

## Data Availability

The full dataset generated and/or analysed during the current study is not publicly available due to the presence of non-anonymised high-resolution medical data. However, we provide an anonymised sample dataset from a single patient to facilitate testing. After installing the SEEG Contact Detector extension, this dataset can be accessed via the Sample Data module in 3D Slicer. Alternatively, it can be downloaded directly from: https://github.com/EpiReC-ISARG/SlicerSEEGContactDetector/releases/tag/v0.1.0

## 5 Availability and requirements

### 5.1 Project name

SEEG Contact Detector.

### 5.2 Project home page

The SEEG Contact Detector extension home page is at: https://epirec-isarg.github.io/SlicerSEEGContactDetector. The full source code of the extension is openly available at: https://github.com/EpiReC-ISARG/SlicerSEEGContactDetector.

### 5.3 Operating system(s)

Platform independent.

### 5.4 Programming language

Python.

### 5.5 Other requirements

The extension has been developed and tested with 3D Slicer versions 5.8.1 and 5.10.0 (available in Extension Manager) on PC:

- Windows 10 (22H2), CPU Intel i7-8700 3.20 GHz, 32 GB RAM, GPU Intel UHD Graphics 630 (128 MB).
- Linux Ubuntu 24.04.4 LTS, CPU AMD Ryzen 7 2700X, 64 GB RAM, GPU NVIDIA GeForce GTX 1050 Ti 4GB VRAM.
- Windows 11 (25H2), CPU Ryzen 9 5950X, 128 GB RAM, GPU NVIDIA RTX 4060 8GB VRAM.

On the Ubuntu system, processing the patient with the highest number of implanted contacts (288 contacts) required 1 minute and 2 seconds to generate the CT brain mask, followed by an additional 8 seconds to detect all electrode contacts.

Recommended configuration: CPU Intel Core i5 / AMD Ryzen 5, 16 GB RAM, GPU 4GB VRAM (not required).

### 5.6 License

Apache-2.0 license.

### 5.7 Any restrictions to use by non-academics

None.

## List of abbreviations

AC-PC: anterior commissure – posterior commissure
CIOMS: Council for International Organizations of Medical Sciences
CT: computed tomography
EP: entry point
GMM: Gaussian mixture model
GT: ground truth
GUI: graphical user interface
HU: Hounsfield units
MNI: Montreal Neurological Institute
MRI: magnetic resonance imaging
PCA: principal component analysis
RAS: “left to Right, posterior to Anterior, inferior to Superior” world coordinate system
ROI: region of interest
SEEG: stereoelectroencephalography
TP: electrode tip
WHO: World Health Organization

## 7 Declarations

## 7.1 Ethics approval and consent to participate

The study was approved by the institutional ethical committee of Motol University Hospital (2022/06/15-EK-602.24/22 and 2024/06/12-EK-279.42/24) in accordance with the Declaration of Helsinki of the World Medical Association and the International Ethical Guidelines for Biomedical Research Involving Human Subjects, prepared by the Council for International Organizations of Medical Sciences (CIOMS) in collaboration with the World Health Organization (WHO), issued in Geneva, 1993.

## 7.2 Consent for publication

Not applicable.

## 7.6 Authors’ contributions

JS: Software; Validation; Investigation; Formal analysis; Visualisation; Writing - Original Draft. RJ: Conceptualisation; Methodology; Writing - Original Draft; Resources; Investigation; Supervision; Project administration; Funding acquisition. PJ: Resources; Data Curation. AK: Software; Validation; Writing - Review & Editing; Resources. MK: Resources. All authors read and approved the final manuscript.

## 7.7 Acknowledgements

The authors would like to thank the participating clinics of Motol and Homolka University Hospital, their heads and staff for their support and cooperation throughout this study: Department of Neurology - Prof. Petr Marusic, MD, PhD; David Krysl, MD, PhD; Department of Pediatric Neurology - Prof. Pavel Krsek, MD, PhD; Matyas Ebel, MD, PhD; Alena Jahodova, MD, PhD; Anezka Hejbalova (Belohlavkova), MD, PhD. Department of Neurosurgery - Assoc. Prof. Vladimir Benes, MD, PhD; Assoc. Prof. Petr Liby, MD, PhD; Robert Lesko, MD, PhD; Department of Radiology - Prof. Lukas Lambert, MD, PhD, MBA.

Access to CESNET storage facilities provided by the project “e-INFRA CZ” under the programme “Projects of Large Research, Development, and Innovations Infrastructures” LM2023054), is greatly acknowledged.

## Footnotes

1 Fiducial placement directly on rendered surfaces is currently not supported when using the experimental VTK Multi-Volume rendering mode. Please use either VTK CPU Ray Casting or VTK GPU Ray Casting.

2 X-ray photon starvation and beam hardening artefacts cause hypointense artefacts with extreme negative HU, usually occurring inside or between the metallic contacts and distorting the result of correlation. Therefore, a uniform filter with a kernel size corresponding to the electrode diameter is applied to the original CT volume. For voxels with intensity below the metal threshold in the original CT, the filtered values are used. This operation replaces hypointense voxels while preserving the high-intensity metal signal of the electrode contacts. The model correlation is performed with filtered CT.

3 The precise alignment to the anterior commissure - posterior commissure (AC-PC) plane was not performed; the orientation was used from the MRI set by an operator.

